# Multicenter cohort study of multisystem inflammatory syndrome in children (MIS-C)

**DOI:** 10.1101/2021.05.14.21257058

**Authors:** Joanna Merckx, Suzette Cooke, Tala El Tal, Ronald M. Laxer, Ari Bitnun, Shaun K. Morris, E. Ann Yeh, Carmen Yea, Peter Gill, Jesse Papenburg, Marie-Astrid Lefebvre, Rolando Ulloa-Gutierrez, Helena Brenes-Chacon, Adriana Yock-Corrales, Gabriela Ivankovich-Escoto, Alejandra Soriano-Fallas, Marcela Hernandez-de Mezerville, Tammie Dewan, Lea Restivo, Alireza Nateghian, Behzad Haghighi Aski, Ali Manafi, Rachel Dwilow, Jared Bullard, Alison Lopez, Manish Sadarangani, Ashley Roberts, Michelle Barton, Dara Petel, Nicole Le Saux, Jennifer Bowes, Rupeena Purewal, Janell Lautermilch, Sarah Tehseen, Ann Bayliss, Jacqueline K. Wong, Kirk Leifso, Cheryl Foo, Joan Robinson, on behalf of the Paediatric Investigators Collaborative Network on Infections in Canada (PICNIC)

## Abstract

**BACKGROUND:** SARS-CoV-2 infection can lead to multisystem inflammatory syndrome in children (MIS-C). We investigated risk factors for severe disease and explored changes in severity over time.

**METHODS:** Children up to 17 years of age admitted March 1, 2020 through March 7^th^, 2021 to 15 hospitals in Canada, Iran and Costa Rica with confirmed or probable MIS-C were included. Descriptive analysis and comparison by diagnostic criteria, country, and admission date was performed. Adjusted absolute average risks (AR) and risk differences (RD) were estimated for characteristics associated with ICU admission or cardiac involvement.

**RESULTS:** Of 232 cases (106 confirmed) with median age 5.8 years, 56% were male, and 22% had comorbidities. ICU admission occurred in 73 (31%) but none died. Median length of stay was 6 days (inter-quartile range 4-9). Children 6 to 12 years old had the highest AR for ICU admission (44%; 95% confidence interval [CI] 34-53). Initial ferritin greater than 500 mcg/L was associated with ICU admission. When comparing cases admitted up to October 31, 2020 to those admitted later, the AR for ICU admission increased from 25% (CI 17-33) to 37% (CI 29-46) and for cardiac involvement from 44% (CI 35-53) to 75% (CI 66-84). Risk estimates for ICU admission in the Canadian cohort demonstrated a higher risk in December 2020-March 2021 compared to March-May 2020 (RD 25%; 95%CI 7-44).

**INTERPRETATION:** MIS-C occurred primarily in previously well children. Illness severity appeared to increase over time. Despite a high ICU admission incidence, most children were discharged within one week.

## Introduction

Multisystem inflammatory syndrome in children (MIS-C) was first recognized in April 2020 (1) and manifests as immune dysregulation following symptomatic or asymptomatic SARS-CoV-2 infection (2). Clinical manifestations overlap with toxic shock syndrome and Kawasaki disease (3). There are no pathognomonic features. Thus, the diagnostic criteria of the Royal College of Paediatrics and Child Health (RCPCH), the Centers for Disease Control (CDC) and the World Health Organization (WHO) differ (3).

The primary objective was to assess factors associated with increased risk for ICU admission or cardiac involvement. We also evaluated changes in severity over time as an exploratory analysis with the hypothesis being that severity increased. MIS-C case series originate primarily from high income countries (4-7) with series from middle income countries just starting to appear (8, 9). A secondary objective was to describe and compare the clinical features and outcome of hospitalized MIS-C cases in a high income country (Canada) with two middle income countries (Costa Rica and Iran) (10).

## Methods

Investigators from 15 pediatric hospitals (13 in Canada and one each in San José, Costa Rica and Tehran, Iran) entered consecutive children up to 17 years of age admitted March 1, 2020 through March 7, 2021 who fulfilled the WHO criteria for MIS-C (11): fever for minimum 3 days, laboratory evidence of inflammation, illness involving two or more systems with no other obvious microbial cause of inflammation, and “evidence of SARS-CoV-2 infection (RT-PCR, antigen test or serology positive), or likely contact with patients with COVID-19” (11). Given that asymptomatic individuals can transmit SARS-CoV-2 (12), any child residing in a community with ongoing SARS-CoV-2 activity was considered to have “likely contact”. In addition, we included patients who otherwise fulfilled WHO criteria but had fever for less than 3 days if they received corticosteroids and/or intravenous immunoglobulin (IVIG) as treatment for MIS-C prior to the third day of fever. Individuals with a likely alternative diagnosis were excluded. Cases were identified by i) screening admission lists, ii) review of admitted children with positive testing for SARS-CoV-2, and iii) communication with the clinical services likely to admit or be consulted on MIS-C cases.

Study data were collected and managed using REDCap electronic data capture tools hosted at the University of Alberta. Ethics approval was obtained initially at the University of Alberta (Pro00099426) and then from all participating sites. Data were collected by chart review on primary reason for admission, demographics, comorbidities, clinical presentation and course, coinfections, treatments and complications.

The STrengthening the Reporting of OBservational studies in Epidemiology (STROBE) guidelines were followed (13).

### Definitions

Cases were considered *confirmed* if in addition to fulfillment of MIS-C clinical criteria, SARS-CoV-2 from any patient specimen or antibodies to SARS-CoV-2 were detected. Cases were considered *probable* if they fulfilled clinical criteria but SARS-CoV-2 tests were negative or not performed. Other definitions are shown in Table 1.

**Table 1.** Study definitions.

| Clinical manifestation | Definition |
| --- | --- |
| Critical disease | Admission to an intensive care unit with use of vasopressors or non-invasive or mechanical ventilation |
| Viral co-infection<sup>1</sup> | Laboratory detection of any virus concurrent with SARS-CoV-2 infection |
| Bacterial co-infection<sup>1</sup> | Laboratory detection of one or more bacteria concurrent with SARS-CoV-2 infection and treated with antibiotics by the attending physician |
| Mucocutaneous involvement | Rash, conjunctivitis, strawberry tongue, red cracked lips and/or redness or swelling of extremities. |
| Gastrointestinal involvement | Vomiting, diarrhea and/or abdominal pain |
| Cardiac involvement | Echocardiographic evidence of pericarditis, valvulitis, coronary vessel abnormalities, or biochemical evidence with troponin >0.5 pg/ml or NT-proB-type Natriuretic Peptide (BNP) >100 pg/ml (11). |
| Acute kidney injury | Serum creatinine above 140 umol/L for children up to 7 days of age, 55 umol/L for children 7 to 364 days of age, 100 umol/L for children 1 to 12 years of age and 140 umol/L for those 13 to 17 years of age (excluding children with chronic renal disease) (14). |
| Hepatitis | Alanine aminotransferase (ALT) >40 IU/L and/or aspartate aminotransferase (AST) > 50 IU/L (15). |
| Neurological complications | Worsening or new onset of seizure, encephalopathy, dystonia, chorea, athetosis, hemiparesis and/or abnormal cerebrospinal fluid cell count for age (>15 X 10<sup>9</sup>/L if less than 30 days of age and >5 X 10<sup>9</sup>/L if older) |
| Clinical hematological complications | Disseminated intravascular coagulation, thrombosis and/or bleeding. |

<sup>1</sup> Investigations were at the discretion of the treating clinician.

### Data analysis

Baseline and demographic characteristics were summarized using descriptive statistics comparing demographic, clinical, laboratory and treatment data as well as outcomes to identify differences between confirmed and probable cases and between the participating countries. Categorical data were compared using chi square or Fisher exact test and continuous data compared using Kruskal-Wallis for medians. Multivariable logistic or linear regression analysis, as appropriate, was used to explore factors associated with length of hospital stay (LOS), ICU admission and cardiac involvement. As criteria for ICU admission can vary by site, the outcome “critical disease” (defined as ICU admission with use of vasopressors or non-invasive or mechanical ventilation) was assessed in a sensitivity analysis. Exposures of interest included sex, age, country, serum laboratory variables including initial ferritin (16), initial white blood cell and platelet count and organ system involvement on admission and are assessed in separate models. Outcomes were compared for two time cohorts – March 1-October 31, 2020 and November 1, 2020-March 7, 2021 and analysed as exposures in separate models. These dates were chosen arbitrarily prior to data analysis to divide the cases into approximately equal groups. It was not possible to analyze cases by epidemic wave as distinct waves did not occur in Costa Rica. Since it may inform practice, data from Canada were analysed separately for risk factors for ICU admission and cardiac involvement, dividing cases into 3-month periods starting March 2020 (with the 3 patients admitted March 2021 added to the fourth quarter).

Results of regression models are reported as adjusted predicted probabilities of the outcome (absolute risks (AR) and risk differences (RD)) representing the average marginal effect across the total study population (17), with a RD not including the null (0%) in its confidence interval (CI) being considered as statistically significant. A directed acyclic graph was made to inform the assessment of potential confounders, mediators and colliders. Model fit was assessed using the area under the curve for logistic and R-squared for linear regression. No adjustments were made for multiple comparisons. Missing data in the included variables were excluded from the analysis. Data were analyzed using STATA 13 (StataCorp. College Station, TX).

## Results

### Demographics

There were 232 MIS-C cases (106 confirmed; 126 probable) (Figure 1). Figure 2 shows the timing of cases. The median age was 5.8 years (IQR 3.0-9.5) (Table 2). Two infants were younger than 90 days (42 and 71 days). Fifty (22%) had one or more comorbidities.

**Table 2.**
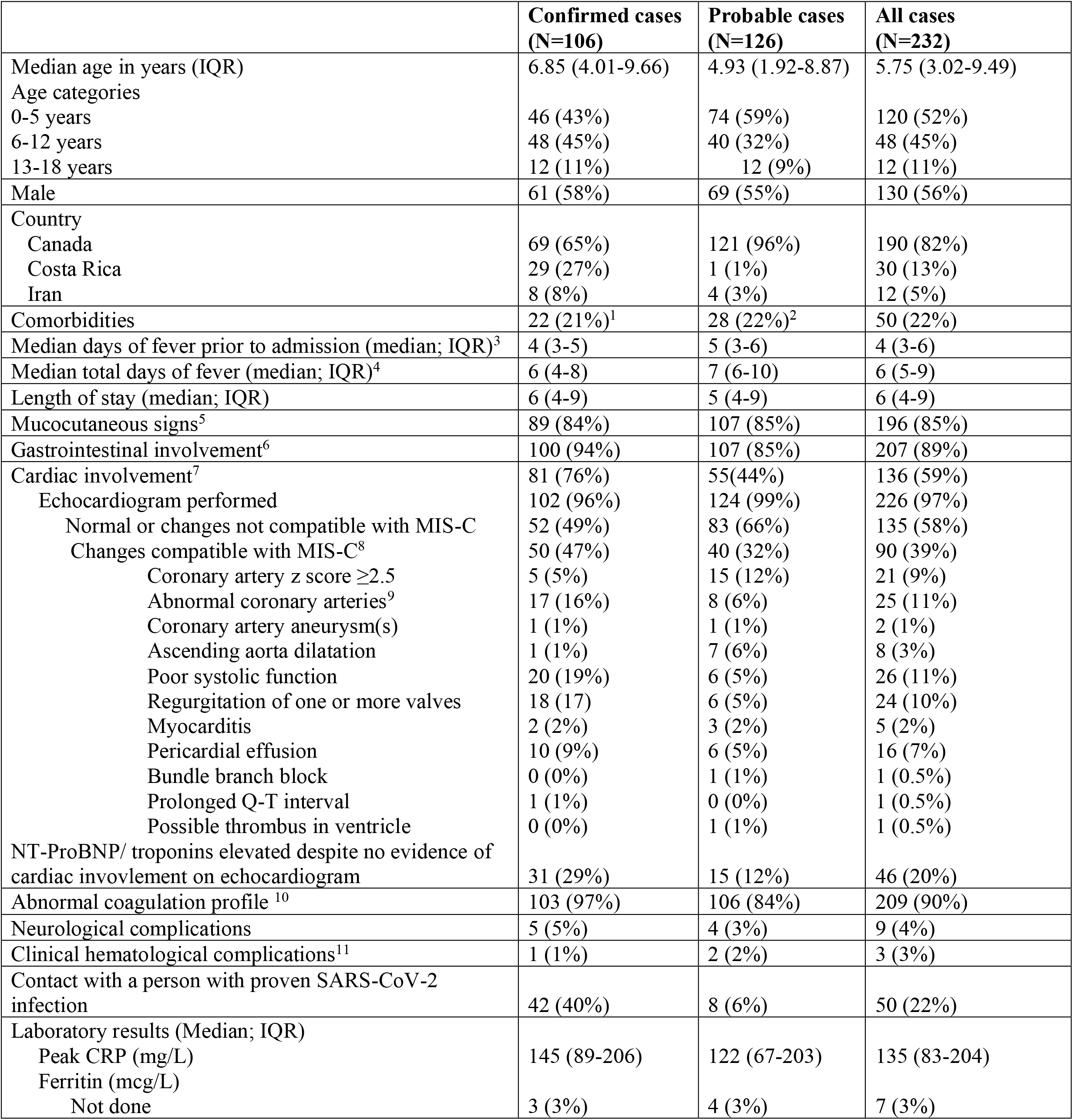
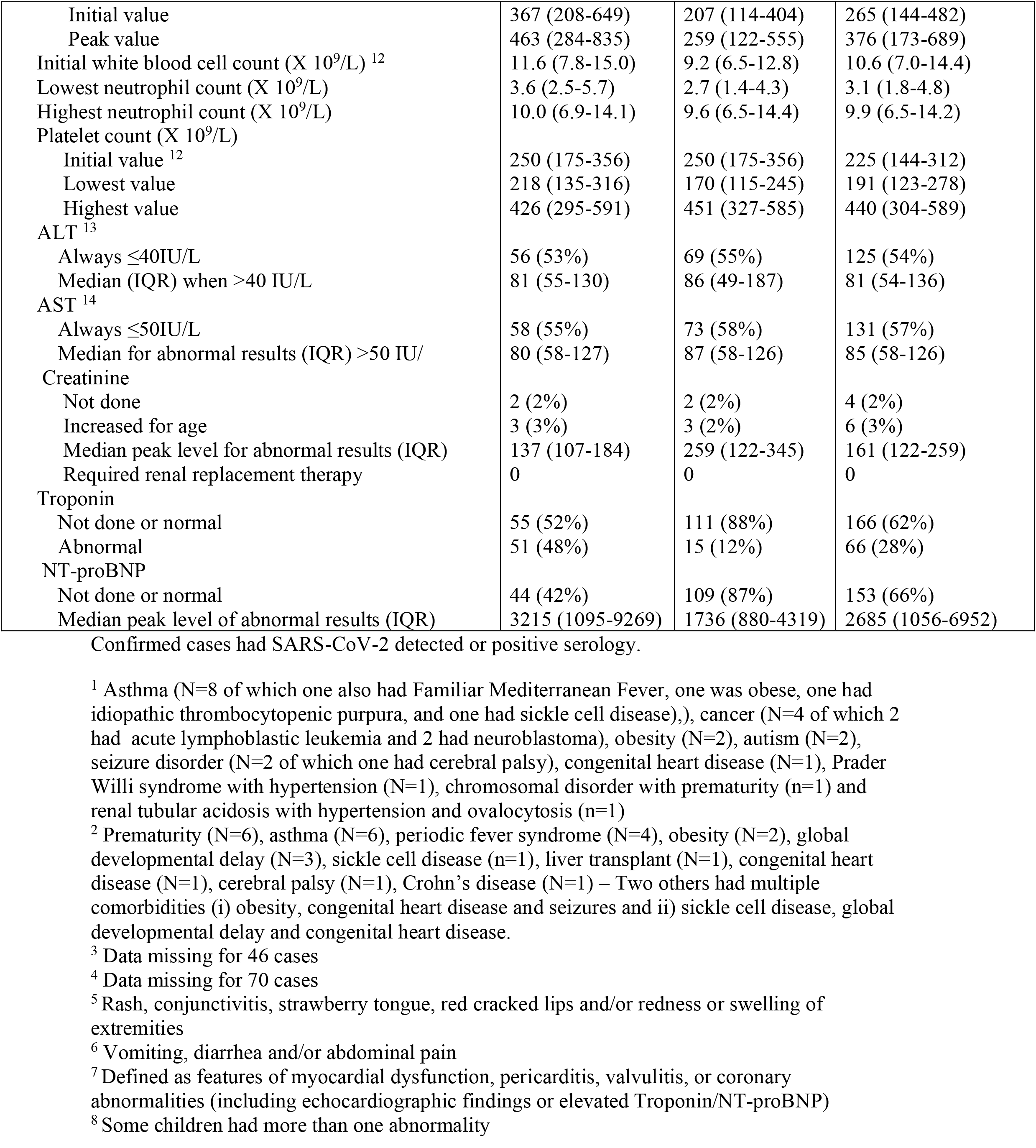
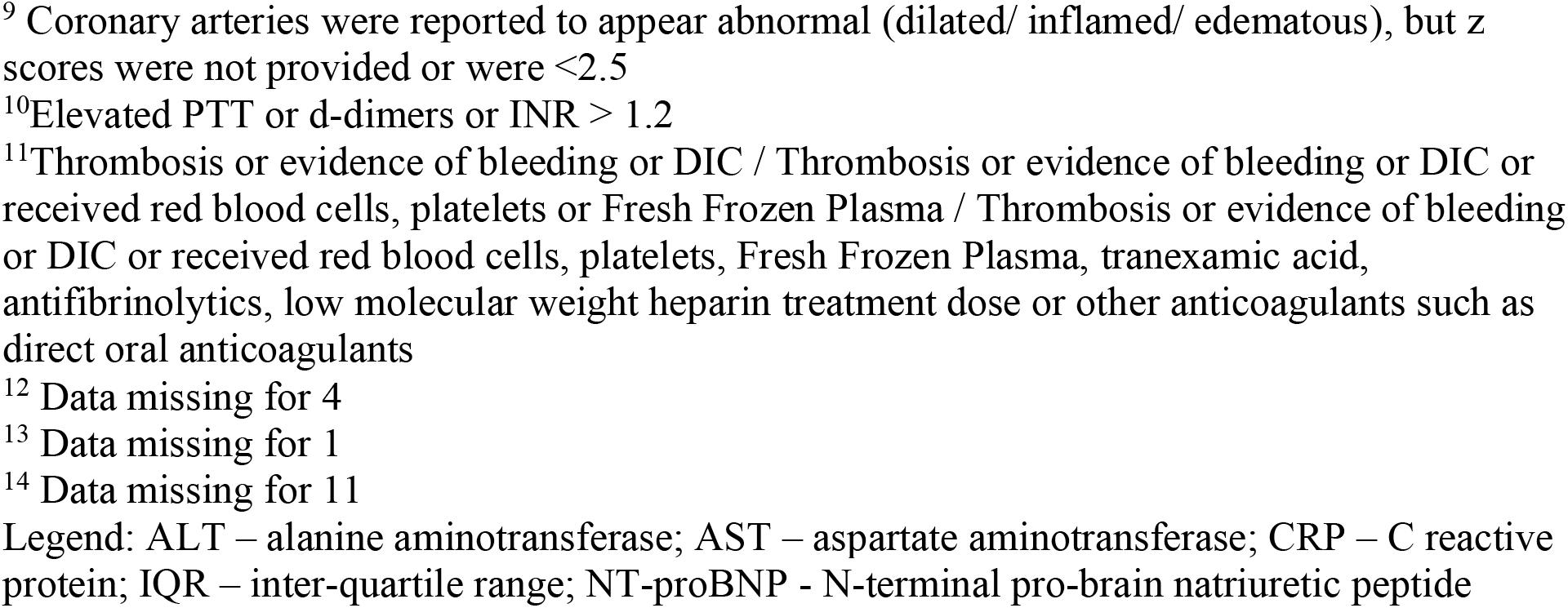
Characteristics of 232 cases of MIS-C.

**Figure 1.**
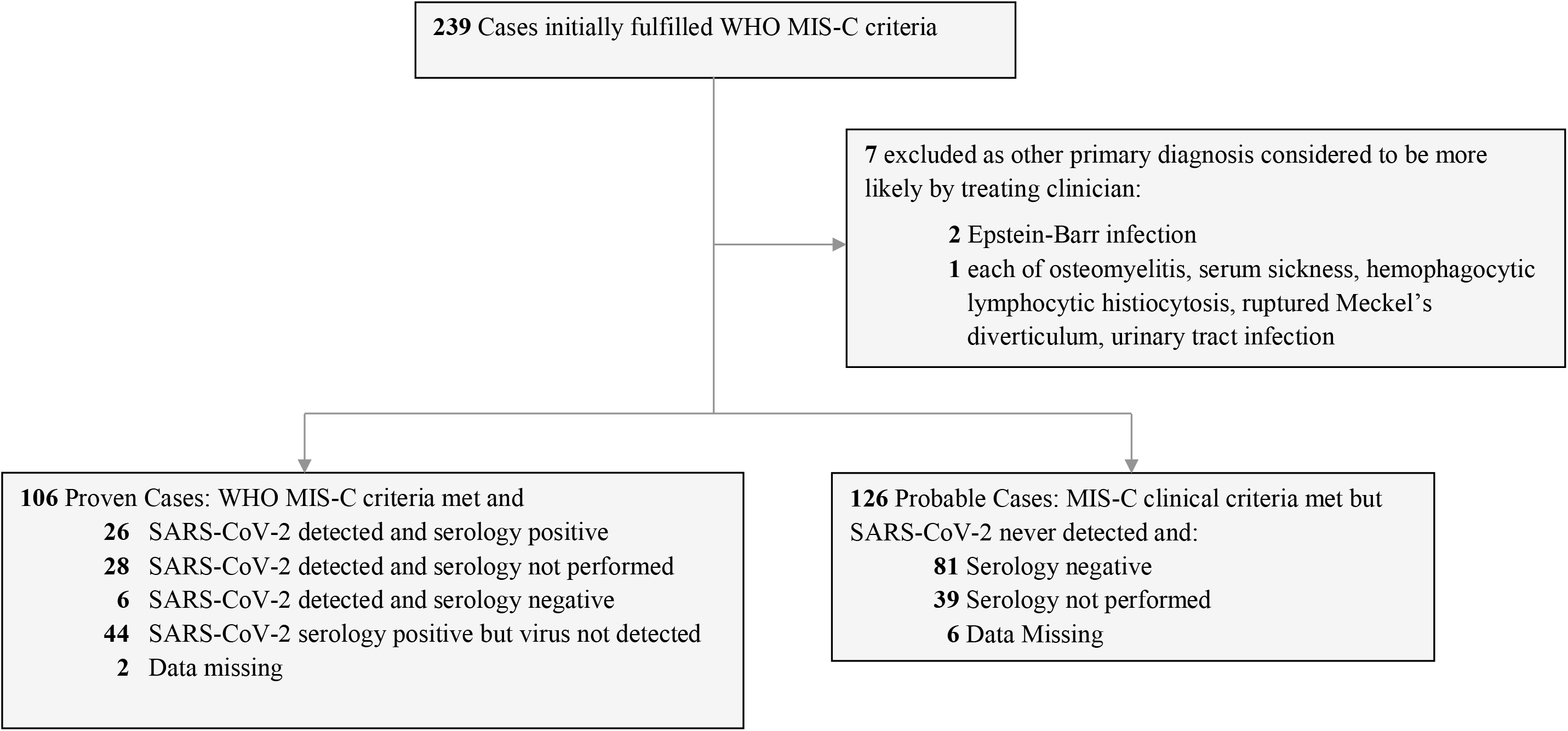
Eligibility flowchart of hospitalized patients with MIS-C.

**Figure 2:**
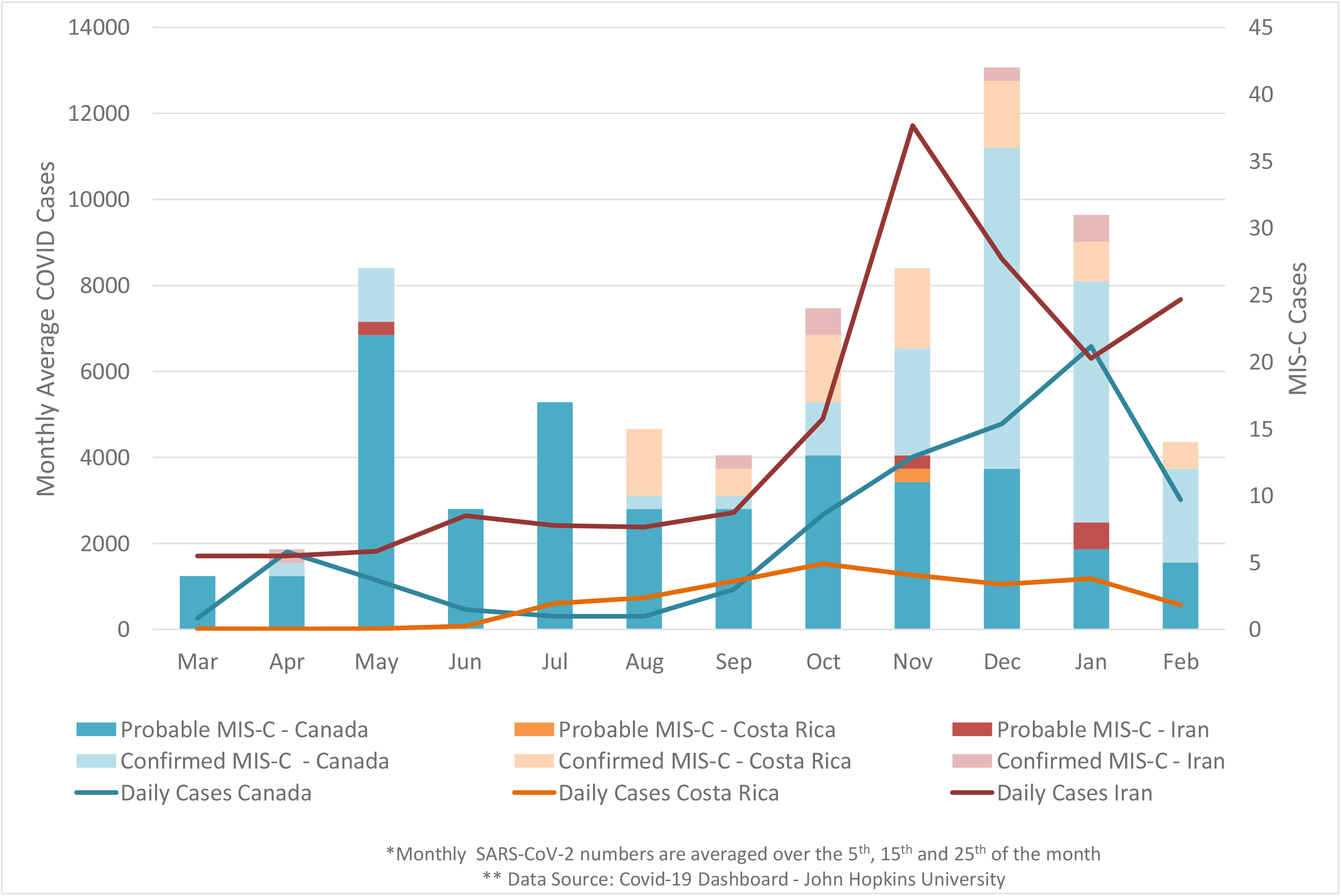
Timing of 232 probable and confirmed cases of MIS-C in Canada, Costa Rica and Iran.

### Clinical features

Almost all (89%) had gastro-intestinal involvement (mainly abdominal pain), and mucocutaneous findings (85%) (Table 2). Confirmed cases had a median total duration of fever of 6 days (IQR 4-8) compared to 7 days (IQR 6-10) for probable cases.

### Coinfections

Bacterial coinfections were documented in 12 cases (5%) (4 had pharyngitis, 3 had colitis and 5 had urinary tract infections) while 142 (61%) received antibiotics, primarily as empiric therapy for sepsis. Viral coinfections were documented in 9 cases (rhinovirus (N=4), rhinovirus/enterovirus (N=3), rhinovirus/enterovirus plus parainfluenza (N=1) and adenovirus (N=1).

### Complications attributed to MIS-C

Cardiac involvement was common among cases (N=136, 59%) including 76% of confirmed and 44% of probable cases (Table 2). Six (3%) had acute kidney injury; none required renal replacement therapy. One hundred twenty (52%) had hepatitis; none progressed to liver failure.

Most cases (N=209; 90%) had abnormalities in one or more laboratory coagulation parameters yet only three patients (1.3%) developed clinical hematological manifestations. One with confirmed MIS-C had extensive venous thrombosis involving the internal jugular vein, a lower extremity and lungs. One with probable MIS-C developed pulmonary emboli and melena and a second had hematemesis with disseminated intravascular coagulation.

Other complications in confirmed cases included removal of a normal appendix (N=3, including one 4 days prior to the MIS-C admission), anterior uveitis (N=2), reversible encephalopathy (N=2) and one case each of ileitis requiring total parenteral nutrition, mesenteric adenitis, and orchitis. Complications in probable cases included: aseptic meningitis (N=5; none had a virus detected), removal of a normal appendix (N=2) and one case each of ascites, pleural effusion, renal infarction, cholecystitis, orchitis and raised intracranial pressure. One confirmed and three probable cases were diagnosed with macrophage activation syndrome by the treating clinicians.

### Treatment

IVIG was given to 194 patients (84%), corticosteroids to 126 (54%) and anakinra to 10 (4%) while 24 (10%) received none of these treatments (Table 3).

**Table 3.** Characteristics of 232 cases of MIS-C, according to country of admission.

|  | **Canada**<br>**(N=190)** | **Costa Rica**<br>**(N=30)** | **Iran**<br>**(N=12)** |
| --- | --- | --- | --- |
| Median age (IQR) | 6.23 (2.80-9.82) | 6.08 (3.93-8.34) | 3.43 (2.03-4.81) |
| Sex - Male | 108 (57%) | 17 (57%) | 5 (42%) |
| Median days of fever prior to admission (IQR) | 5 (3-6) | 4 (3-6) | 3.5 (3-5) |
| Median total duration of fever in days (IQR) | 6 (5-8) | 5 (4-8) | 8.5 (7-10) |
| Mucocutaneous signs | 159 (84%) | 29 (97%) | 8 (67%) |
| Gastrointestinal involvement | 165 (87%) | 30 (100%) | 12 (100%) |
| Median length of stay in days (IQR) | 5 (4-8) | 7 (5-9) | 9.5 (8-11) |
| Highest level of care required |  |  |  |
| Ward admission: | 128 (67%) | 26 (87%) | 5 (42%) |
| No supplemental oxygen | 118 | 21 | 4 |
| Supplemental oxygen | 10 | 5 | 1 |
| ICU admission: | 62 (33%) | 4 (13%) | 7 (58%) |
| Observation +/- supplemental oxygen | 17 | 1 | 4 |
| Required vasopressors | 33 | 0 | 2 |
| Required non-invasive ventilation | 4 | 0 | 0 |
| Required mechanical ventilation | 4 | 3 | 1 |
| Admitted for indications other than MIS-C | 4 | 0 | 0 |
| Cardiac involvement | 105 (55%) | 26 (87%) | 5 (42%) |
| Treatment<sup>1</sup> |  |  |  |
| IVIG | 153 (81%) | 29 (97%) | 12 (100%) |
| Corticosteroids | 104 (55%)<sup>2</sup> | 14 (47%)<sup>3</sup> | 8 (75%)<sup>4</sup> |
| Anakinra | 10 (5%) | 0 (0%) | 0 (0%) |
| None of the above | 23 (12%) | 1 (3%) | 0 (0%) |

Legend: IQR – inter-quartile range; IVIG – intravenous immunoglobulin
<sup>1</sup> Many patients received a combination of these treatments.
<sup>2</sup> 66 children were given 1 to 3 days of pulse methylprednisolone (>10 mg/kg/day)
<sup>3</sup> No children were given pulse methylprednisolone (>10 mg/kg/day)
<sup>4</sup> One child was given pulse methylprednisolone (>10 mg/kg/day)

### Outcomes

Table 3 compares cases in Canada (N=190; 82%) to Costa Rica (N=30; 13%) and Iran (N=12; 5%). There was a trend towards a longer duration of fever and a higher incidence of ICU admission in Iran. The median length of hospital stay (LOS) for the 232 cases was 6 days, with 69% being discharged by day 7 and 85% by day 10. Seventy-three (31%) required ICU admission, of which 47 (64%) required vasopressors and/or mechanical or non-invasive ventilation. None required extracorporeal life support or died. Table 4 provides comparison between MIS-C patients admitted by October 31, 2020 (N=120) versus patients admitted later (N=112). The earlier cohort had 23 patients requiring ICU admission (19%) versus 50 in the later cohort (45%) (p<0.001).

**Table 4.** Comparison of cases of MIS-C according to time of admission.

|  | *Admitted Feb 1 to Oct 31, 2020* |  |  | *Admitted Nov 1, 2020 – Mar 7, 2021* |  |  |
| --- | --- | --- | --- | --- | --- | --- |
|  | **Confirmed**<br>**(N=28)** | **Probable**<br>**(N=92)** | **All**<br>**(N=120)** | **Confirmed**<br>**(N=78)** | **Probable**<br>**(N=34)** | **All**<br>**(N=112)** |
| Country |  |  |  |  |  |  |
| Canada | 11(39%) | 91 (99%) | 102 (85%) | 58 (74%) | 30 (88%) | 88(79%) |
| Costa Rica | 13 (47%) | 0 (0%) | 13 (11%) | 16 (21%) | 1 (3%) | 17 (15%) |
| Iran | 4 (14%) | 1 (1%) | 5 (4%) | 4 (5%) | 3 (9%) | 7 (6%) |
| Median days of fever prior to admission (IQR) | 5 (3-6) | 5 (3-6) | 5 (3-6) | 4 (3-5) | 5 (3-6) | 4 (3-6) |
| Median total duration of fever in days (IQR) | 7 (6-9) | 6.5 (5-9.5) | 7(5-9) | 5.5 (4-8) | 6 (6-10) | 6 (4-8) |
| Median duration of admission in days (IQR) | 8 (6.5-10) | 5 (3-7) | 5 (4-8) | 6 (4-8) | 7 (4-12) | 6 (4-9) |
| Highest level of care |  |  |  |  |  |  |
| Ward admission: | 18 (64%) | 79 (86%) | 97 (81%) | 39 (50%) | 23 (68%) | 62 (55%) |
| No supplemental oxygen | 14 | 76 | 90 | 33 | 20 | 53 |
| Supplemental oxygen | 4 | 3 | 7 | 6 | 3 | 9 |
| ICU admission: | 10 (35%) | 13 (14%) | 23 (19%) | 39 (50%) | 11 (32%) | 50 (45%) |
| Observation +/- supplemental oxygen | 5 | 2 | 7 | 11 | 4 | 15 |
| Vasopressors +/- supplemental oxygen | 3 | 6 | 9 | 19 | 6 | 25 |
| Non-invasive ventilation + vasopressors | 0 | 0 | 0 | 3 | 1 | 4 |
| Mechanical ventilation – no vasopressors | 0 | 1 | 1 | 1 | 0 | 1 |
| Mechanical ventilation + vasopressors | 2 | 1 | 3 | 4 | 0 | 4 |
| Admission not for MIS-C | 0 | 3 | 3 | 1 | 0 | 1 |
| Treatment<sup>1</sup> |  |  |  |  |  |  |
| IVIg | 25 (89%) | 69 (75%) | 94 (78%) | 68 (87%) | 32 (94%) | 100 (89%) |
| Corticosteroids | 12 (43%) | 37 (40%) | 49 (41%) | 57 (73%) | 20 (59%) | 77 (69%) |
| Anakinra | 0 (0%) | 3 (3%) | 3 (3%) | 3 (4%) | 4 (12%) | 7 (6%) |
| None of the above | 2 (7%) | 17 (18%) | 19 (16%) | 4 (5%) | 1 (3%) | 5 (4%) |

<sup>1</sup> Many patients received a combination of these treatments.

### Factors associated with ICU admission

Adjusted RD’s for ICU admission and for cardiac involvement are presented in Figures 3 and 4 and Supplementary Tables 1 and 2. Children 6 to 12 years old had an AR of 44% (95% confidence interval [CI] 34-53) for ICU admission, similar to children 13 to 17 years old (AR 46%, CI 28-64), but significantly higher than children 0 to 5 years old (AR 18%, CI 12-25; RD 25, CI 14-37). An initial ferritin greater than 500 mcg/L was associated with an 18 percentage points higher AR for ICU admission (95% CI 6-31) versus a lower value. Children in Costa Rica had a lower probability, and in Iran, a higher probability for ICU admission compared to those in Canada. As compared to earlier cases, those identified after October 31, 2020 were associated with a non-statistically significant increased risk for ICU admission (AR 37%, CI 29-46; RD 12, CI -0.3-25) but a significantly increased risk for cardiac involvement (AR 75%, CI 66%-84%; RD 31%, CI 17-44). The sensitivity analysis for critical disease confirmed the findings for ICU admission, with the addition of a higher risk for critical disease if presenting with mucocutaneous symptoms (RD 14%, CI 5-23) and also a RD of 13 (CI 2-25) between later versus earlier admission (Supplementary Table 3 and Supplementary Figure 1). Risk estimates for ICU admission and cardiac involvement in the Canadian cohort demonstrated a higher risk for ICU admission in the last quarter compared to March-May 2020 (RD 25%; CI 7-44) (Supplementary Figures 2 and 3). Figure 5 shows the directed acrylic graph.

**Figure 3:**
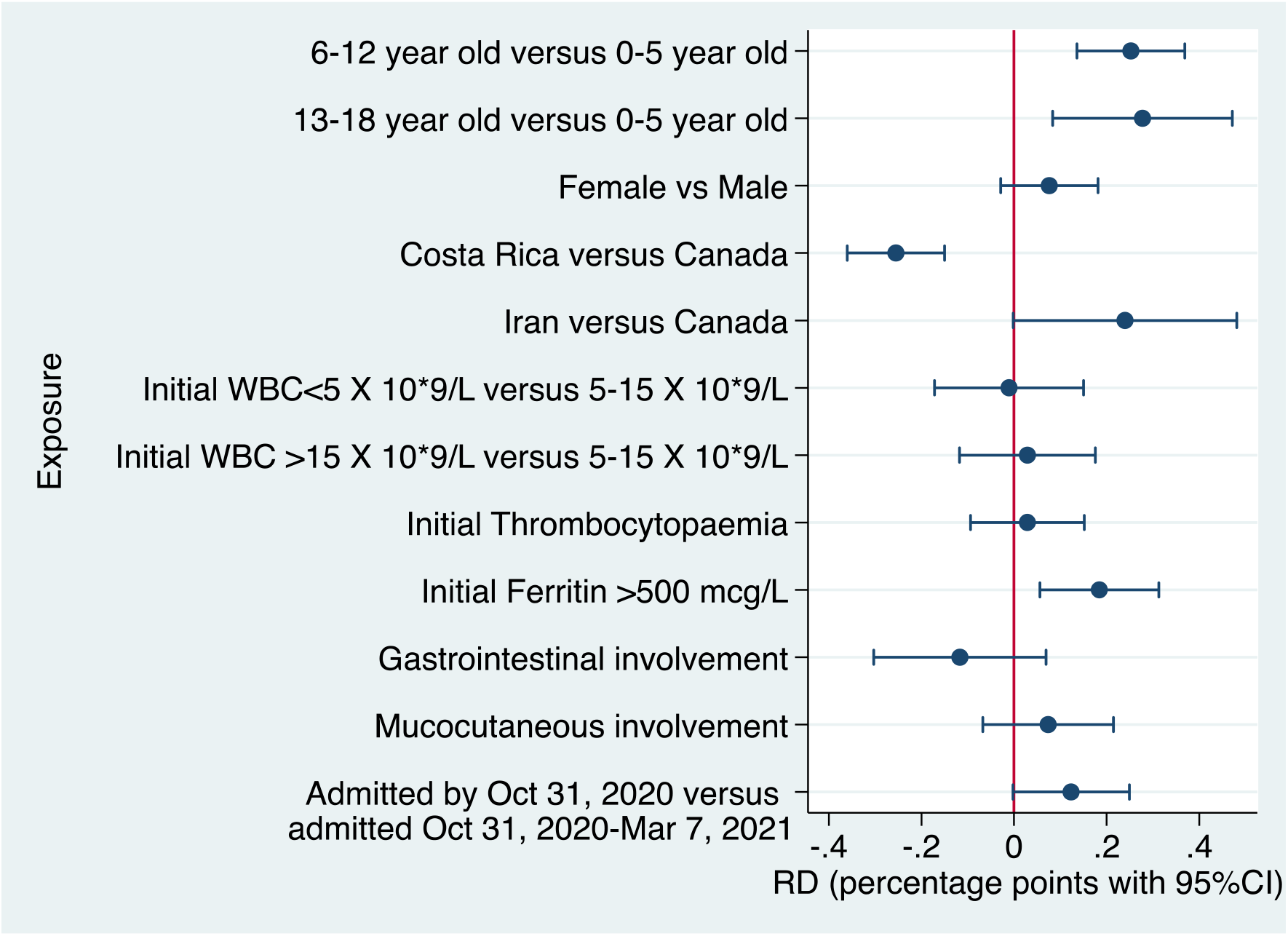
Adjusted Risk Differences for intensive care unit admission. Legend: Average marginal effects are presented with adjusted RDs from multivariable logistic regressions. The reference used for RDs are absence of the symptom or the reported category. Models are adjusted for sex, age, country, presence of comorbidity, coinfection, treatment, MIS-C laboratory confirmation and admission time.

**Figure 4:**
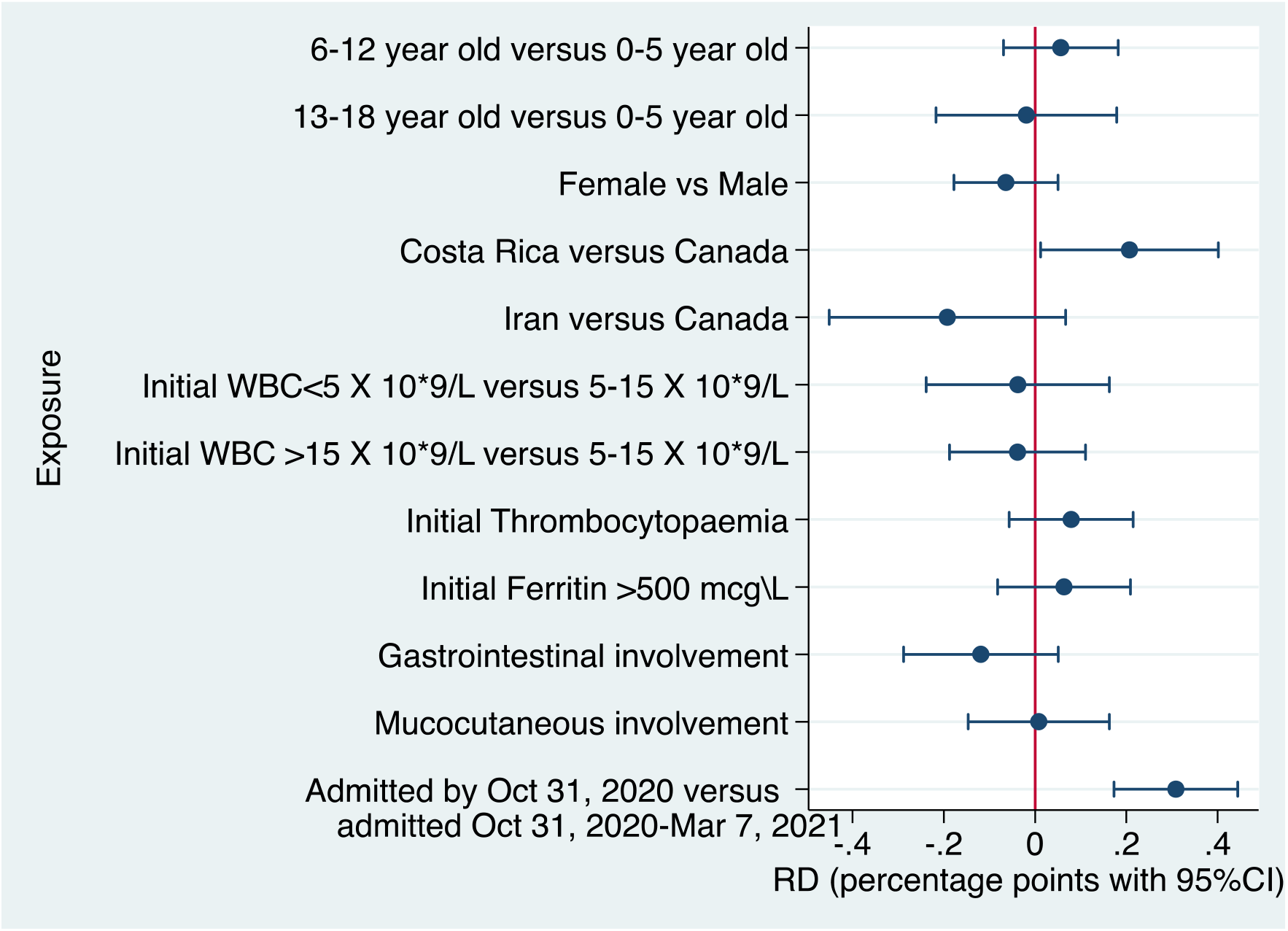
Adjusted Risk Differences for cardiac involvement. Legend: Average marginal effects are presented with adjusted RDs from multivariable logistic regressions. The reference used for RDs are absence of the symptom or the reported category. Models are adjusted for sex, age, country, presence of comorbidity, coinfection, MIS-C laboratory confirmation and admission time.

**Figure 5:**
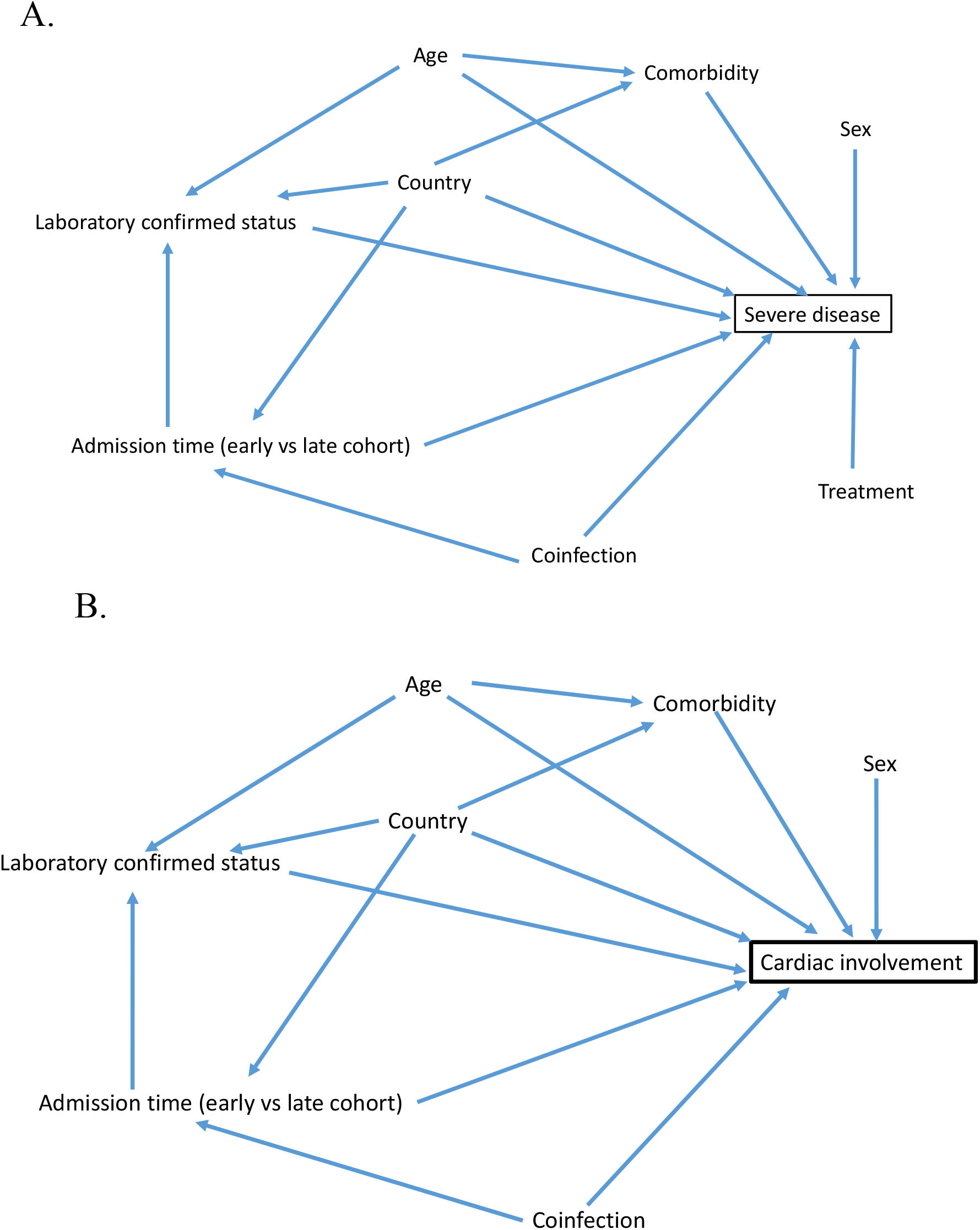
Directed acyclic graph showing risk factors for A. ICU admission and B. cardiac involvement.

## Interpretation

In this multicenter, multinational cohort of children admitted for MIS-C, older age and initial ferritin level greater than 500 mcg/L were associated with ICU admission. Exploratory analyses demonstrated that the adjusted AR for cardiac involvement increased in the second period of the study, with a trend towards a higher incidence of ICU admission. For the Canadian cohort, the risk of ICU admission and/or cardiac involvement was highest in patients admitted in the last quarter of the study. Despite nearly one-third of cases requiring ICU care, recovery was typically rapid with 85% of cases discharged within 10 days.

MIS-C diagnostic criteria may evolve over time. One important difference between criteria is the requirement for evidence of SARS-CoV-2 infection (3). In the absence of viral detection or positive serology, CDC criteria require likely exposure within 4 weeks. WHO criteria do not specify the timing of exposure and do not clarify whether exposure must be to a specific person or includes residence in a community with circulating cases (the definition applied in the current study). RCPCH criteria do not require a history of exposure so all our probable cases would have met these criteria. Most of our 126 probable cases lacked a history of contact with a person with proven SARS-CoV-2 infection. However, 24 (19%) required intensive care and 55 (44%) had evidence of cardiac involvement, making it likely that some had MIS-C. Identifying exposure can be difficult as infected contacts may be asymptomatic or never tested. Laboratory confirmation is also complicated as serology is not 100% sensitive (18) and was not available until partway through the study. As more individuals are infected or vaccinated, the diagnostic utility of serology will decline.

As previously reported (19), patients in the current study typically improved rapidly following IVIG. The role for adjunctive glucocorticoids and the optimal glucocorticoid regimen has not been established. It is not known whether corticosteroids could replace IVIG in low-income countries where IVIG is not readily available. Of note, 24 (10%) of our cases resolved without IVIG or corticosteroids.

Risk factors for ICU admission in 1080 children entered in the CDC database up to October 10, 2020 included older age (13 to 20 years [adjusted odds ratio (OR) 2.6] or 6 to 12 years (OR 1.9) as compared to 0 to 5 years (16). Peak inflammatory markers, brain natriuretic peptide (BNP), serum troponin and nadir platelet counts were predictive of ICU admission, but peak and nadir results are of limited practical value compared to admission values. High initial ferritin was associated with ICU admission in the current study with 55% of those with a level above 500 mcg/L requiring ICU admission (versus 24% with a lower level).

The incidence of cardiac involvement increased in the latter half of the study with a trend towards increased ICU admission. However, the severity appeared to be less than in US cohorts where 74% (5) and 58% (4) required ICU care, versus 31% in the current study. Mechanical ventilation was required in 17% in one US cohort (5) versus 4% in the current study. Vasopressors were required in 45% in one US cohort (5) versus 20% in the current study (5). Differences in case ascertainment (CDC versus WHO criteria), patient management and threshold for ICU admission may account for some of the variability.

There is not a clear explanation for the increase in severity over time. The features of severe MIS-C were widely publicized by May 2020 so it seems unlikely that severe cases were missed early in the study period. SARS-CoV-2 is continuously mutating. It is possible that the immune response to circulating variants alters the severity of COVID-19 and MIS-C when compared to wild-type virus.

Complications of MIS-C continue to be recognized. Six of our cases (2%) developed acute kidney injury versus 9 of 539 (1.7%) in a recent MIS-C series (5). All cases in that series required dialysis while none or ours did. Hepatitis occurred in 120 of our cases (52%) versus 19 of 44 (43%) in a series that included one case of liver failure (15). Five of our cases (2%) had removal of a normal appendix. A February 2021 review of the literature described 8 cases of appendectomy of which two had proven appendicitis (20). Two cases had reversible encephalopathy as previously described (21, 22). One case had raised intracranial pressure as described in 4 previous cases (23). Two cases had anterior uveitis, previously described in four children (24-26). Two cases had pulmonary emboli as described in one previous report (27). Two cases had orchitis while one case of epididymitis was reported previously (28). Renal infarction was documented in one case and has been described in adults with acute COVID-19 (29) but we could find no previous reports in MIS-C.

One limitation is the lack of long-term follow-up. It is encouraging that the cases series from the US reported resolution of cardiac aneurysms in 45 of 57 cases (79%) and normalization of left ventricular function in 91% of 172 cases 30 days after the initial echocardiogram (5). Further limitations include that milder cases might not have been admitted or recognized, SARS-CoV-2 serology was not available initially, and race/ethnicity is not reliably recorded in health records in Canada.

The morbidity from MIS-C probably exceeds the morbidity from acute COVID-19 in children but is over-shadowed by the high morbidity and mortality rate of COVID-19 in adults. The short-term prognosis for MIS-C is generally good but there is a great need for population-based studies with controls to establish the incidence, spectrum, long-term prognosis and evolution of pediatric COVID-19 and MIS-C.

## Data Availability

Study data were collected and managed using REDCap electronic data capture tools hosted at the University of Alberta. Contact the corresponding author regarding data availability.

## Figure Legends

**Supplementary Table 1.** Risk factors for ICU admission (defined as intensive care unit admission) in 232 children with MIS-C - adjusted model estimates.

|  | AR %(95%CI) | RD (95%CI) | RR (95%CI) |
| --- | --- | --- | --- |
| **ICU admission** |  |  |  |
| Age 0-5 years | 18.4 (11.6-25.2) | - | - |
| Age 6-12 years | 43.6 (34.4-52.8) | 25.2 (13.6-36.9) | 2.37 (1.34-3.40) |
| Age 13-18 years | 46.2 (28.2-64.1) | 27.7 (8.3-47.2) | 2.51 (1.14-3.88) |
| Male | 28.0 (21.3-34.8) | - | - |
| Female | 35.6 (27.7-43.5) | 7.6 (-2.9-18.1) | 1.27 (0.85-1.69) |
| Canada | 34.7 (28.5-40.9) | - | - |
| Costa-Rica | 9.2 (1.0-17.4) | -25.5 (-36.0-(-15.0)) | 0.26 (0.03-0.51) |
| Iran | 58.7 (28.5-40.9) | 24.0 (2.3-50.9) | 1.69 (0.95-2.43) |
| Initial WBC<5000 X 10<sup>9</sup>/L | 29.2 (14.6-43.9) | -1.1 (-17.2-15.0) | 0.96(0.44-1.49) |
| WBC 500-15,000 X 10<sup>9</sup>/L | 30.3 (24.1-36.6) | - | - |
| WBC >15,000 X 10<sup>9</sup>/L | 33.2 (20.2-46.2) | 2.9 (-11.8-17.5) | 1.10 (0.60-1.59) |
| Initial platelets <150,000 X 10<sup>9</sup>/L | 33.8 (23.6-44.0) | 2.9 (-9.4-15.2) | 1.09 (0.69-1.50) |
| Initial platelets ≥150,000 X 10<sup>9</sup>/L | 30.9 (24.8-37.1) | - | - |
| Initial ferritin >500 mcg/L | 45.1 (33.8-56.4) | 18.4 (5.6-31.3) | 1.69 (1.12-2.26) |
| Gastrointestinal involvement | 30.4 (25.2-35.6) | -11.7 (-30.3-6.9) | 0.72 (0.39-1.05) |
| Mucocutaneous involvement | 32.6 (27.0-38.1) | 7.4 (-6.8-21.5) | 1.29 (0.59-1.99) |
| Admission prior to Nov 1, 2020 | 24.9 (16.8-33.0) | - | - |
| Admission Nov 1,<br>2020-Mar 7, 2021 | 37.2 (28.9-45.6) | 12.3(-0.3-25.0) | 1.50 (0.86-2.13) |

Models are adjusted for sex, age, country, presence of comorbidity, coinfection, MIS-C laboratory confirmation, treatment and admission time.
Average marginal effects are presented with adjusted RDs from multivariable logistic regressions.

**Supplementary Table 2.** Risk factors for cardiac involvement in 232 children with MIS-C - adjusted model estimates.

|  | AR %(95%CI) | RD (95%CI) | RR (95%CI) |
| --- | --- | --- | --- |
| **Cardiac involvement<sup>1</sup>** |  |  |  |
| Age 0-5 years | 56.8 (48.9-64.7) | - | - |
| Age 6-12 years | 62.4 (52.9-71.9) | 5.6 (-7.0-18.2) | 1.10 (0.87-1.33) |
| Age 13-18 years | 54.9 (37.1-72.8) | -1.9 (-21.7-17.9) | 0.97 (0.62-1.31) |
| Male | 61.5 (54.0-68.9) | - | - |
| Female | 55.1 (46.6-63.6) | -6.4 (-17.8-5.0) | 0.90 (0.72-1.07) |
| Canada | 57.4 (51.0-63.9) | - | - |
| Costa-Rica | 78.1 (60.3-95.9) | 20.7 (12.0-40.1) | 1.36 (1.00-1.72) |
| Iran | 38.2 (13.3-63.1) | -19.2 (-45.1-6.7) | 0.67 (0.22-1.11) |
| Initial WBC<5000 X 10<sup>9</sup>/L | 55.8 (37.0-74.5) | -3.8 (-23.8-16.3) | 0.94 (0.60-1.27) |
| WBC 500-15,000 X 10<sup>9</sup>/L | 60.0 (52.6-66.5) | - | - |
| WBC >15,000 X 10<sup>9</sup>/L | 55.7 (42.7-68.7) | -3.8 (-18.7-11.1) | 0.94 (0.69-1.18) |
| Initial platelets <150,000 X 10<sup>9</sup>/L | 64.6 (53.1-76.0) | 7.9 (-5.7-21.5) | 1.14 (0.89-1.39) |
| Initial platelets ≥150,000 X 10<sup>9</sup>/L | 56.7 (50.0-63.3) | - | - |
| Initial ferritin >500 mcg/L | 64.6 (51.8-77.3) | 6.3 (-8.2-20.9) | 1.11 (0.85-1.36) |
| Gastrointestinal involvement | 57.2 (51.2-63.2) | -11.9 (-28.8-5.1) | 0.83 (0.62-1.03) |
| Mucocutaneous involvement | 58.8 (52.6-64.9) | 0.8 (-14.6-16.3) | 1.01 (0.74-1.28) |
| Admission prior to Nov 1, 2020 | 44.1 (34.9-53.3) | - | - |
| Admission Nov 1, 2020-Mar 7, 2021 | 75.0 (66.4-83.6) | 30.9 (17.3-44.4) | 1.70 (1.27-2.13) |

<sup>1</sup> Defined as features of myocardial dysfunction, pericarditis, valvulitis, or coronary abnormalities (including echocardiographic findings or elevated Troponin/NT-proBNP)
Models are adjusted for sex, age, country, presence of comorbidity, coinfection, MIS-C laboratory confirmation and admission time.

**Supplementary Table 3.**
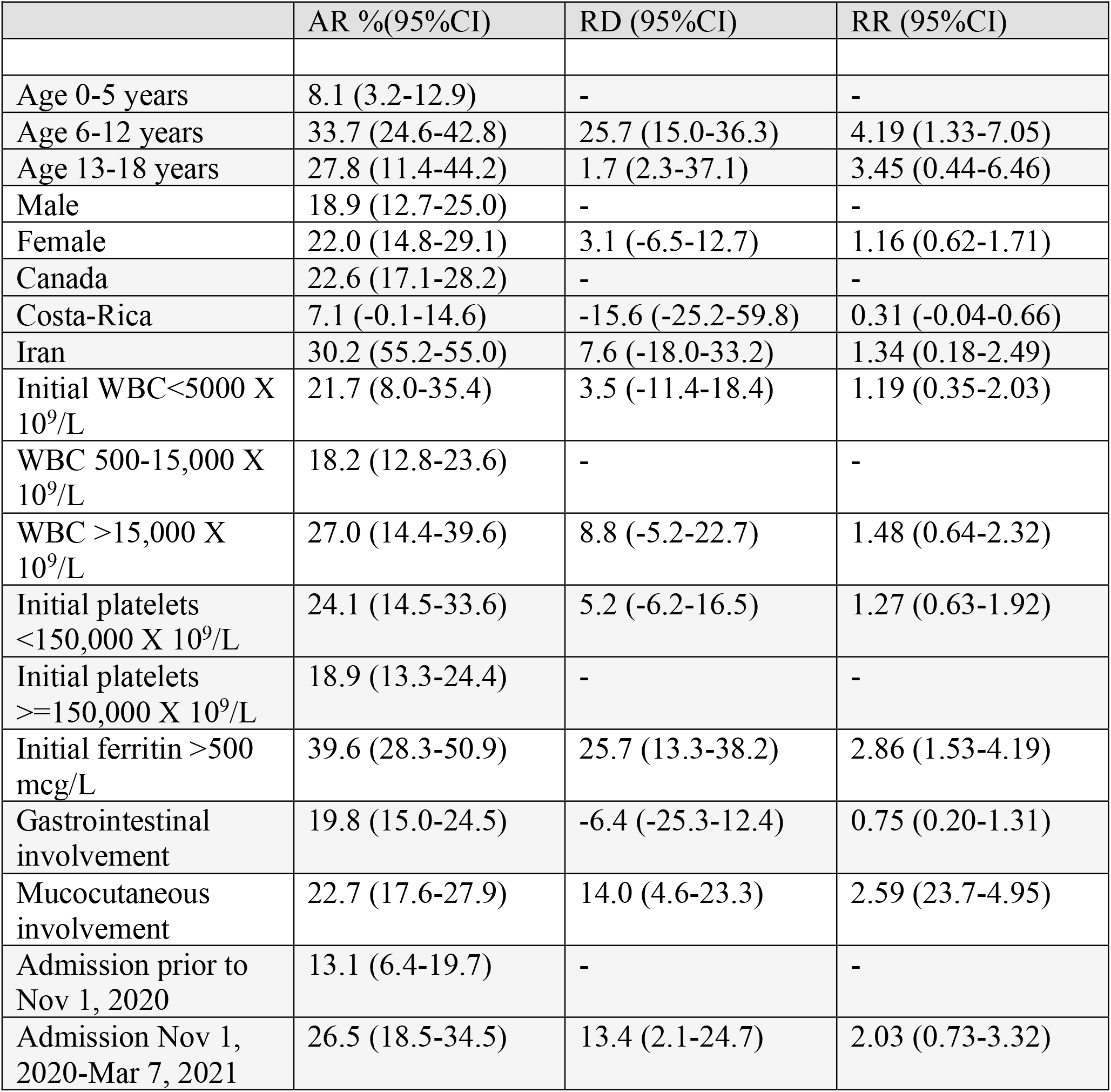
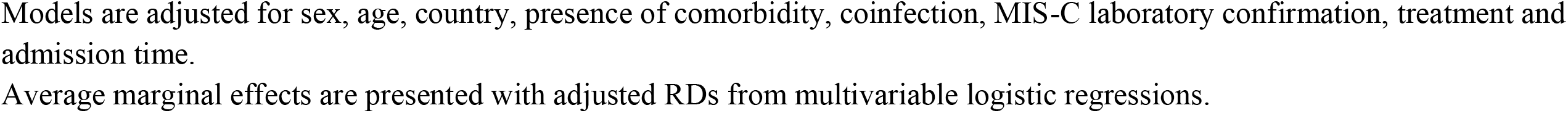
Risk of critical disease (defined as intensive care unit admission with need for vasopressors or ventilation) in 232 children with MIS-C – adjusted model estimates.

**Supplementary Table 4.** Risk of severe MIS-C (defined as intensive care unit admission) in 190 children in Canada by quarter of the year.

|  | AR %(95%CI) | RD (95%CI) | RR (95%CI) |
| --- | --- | --- | --- |
| **ICU admission** |  |  |  |
| Admission March-May 2020 (Q1) | 17.8 (3.8-31.8) | - | - |
| Admission June-August (Q2) | 32.4 (16.4-48.3) | 14.6 (-5.9-35.1) | 1.82 (0.18- 3.47) |
| Admission September-November (Q3) | 24.2 (13.1-35.2) | 6.4 (-11.2-24.1) | 1.36 (0.13- 2.59) |
| Admission Dec 2020-Feb 2021 (Q4) | 43.1 (31.8-54.3) | 25.3 (6.5-44.0) | 2.42 (0.36- 4.50) |

**Supplementary Figure 1.**
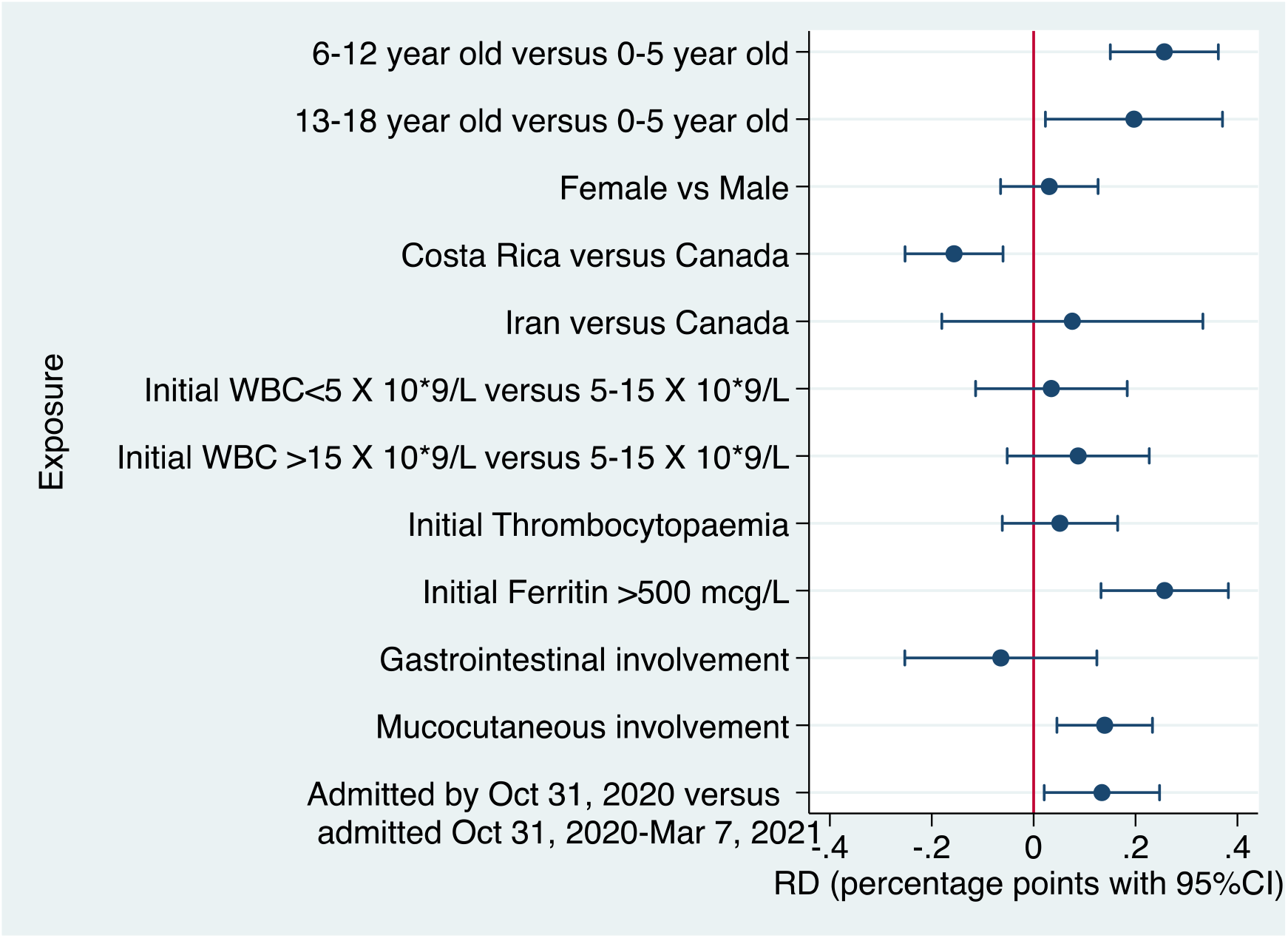
Risk of critical disease (defined as intensive care unit admission with need for vasopressors and/ or non-invasive or mechanical ventilation) in 232 children admitted with MIS-C. Legend: Association exposures on admission and admission on the ICU with the need for vasopressors or ventilation. Average marginal effects are presented with adjusted RDs from multivariable logistic regressions. The reference used for RDs are absence of the symptom or the reported category. Models are adjusted for sex, age, country, presence of comorbidity, coinfection, treatment, MIS-C laboratory confirmation and admission time.

**Supplementary Figure 2.**
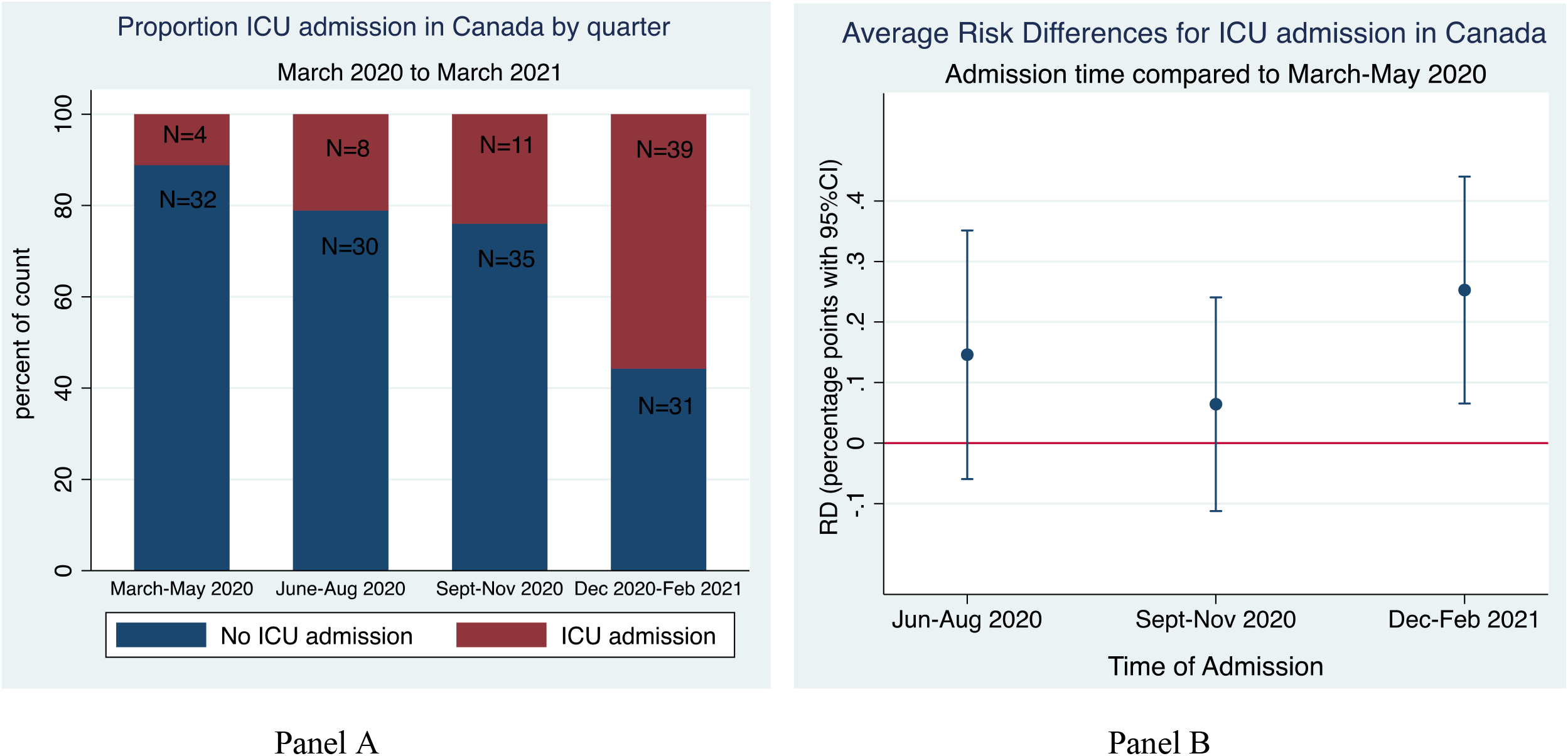
Intensive care unit admission disease in 190 children admitted in Canada with MIS-C by quarter of the year. Panel A: N is the absolute number of cases in the group. The Y-axis reflects the proportion of total cases admitted per quarter. Panel B: Association time of admission and severe disease, defined as need for ICU admission in Canada. Average marginal effects are presented with adjusted RDs from the multivariable logistic regression. The reference used for RDs is admission to the hospital March-May 2020 with a diagnosis of MIS-C. Model is adjusted for sex, age, country, presence of comorbidity, coinfection, treatment, MIS-C laboratory confirmation.

**Supplementary Figure 3.**
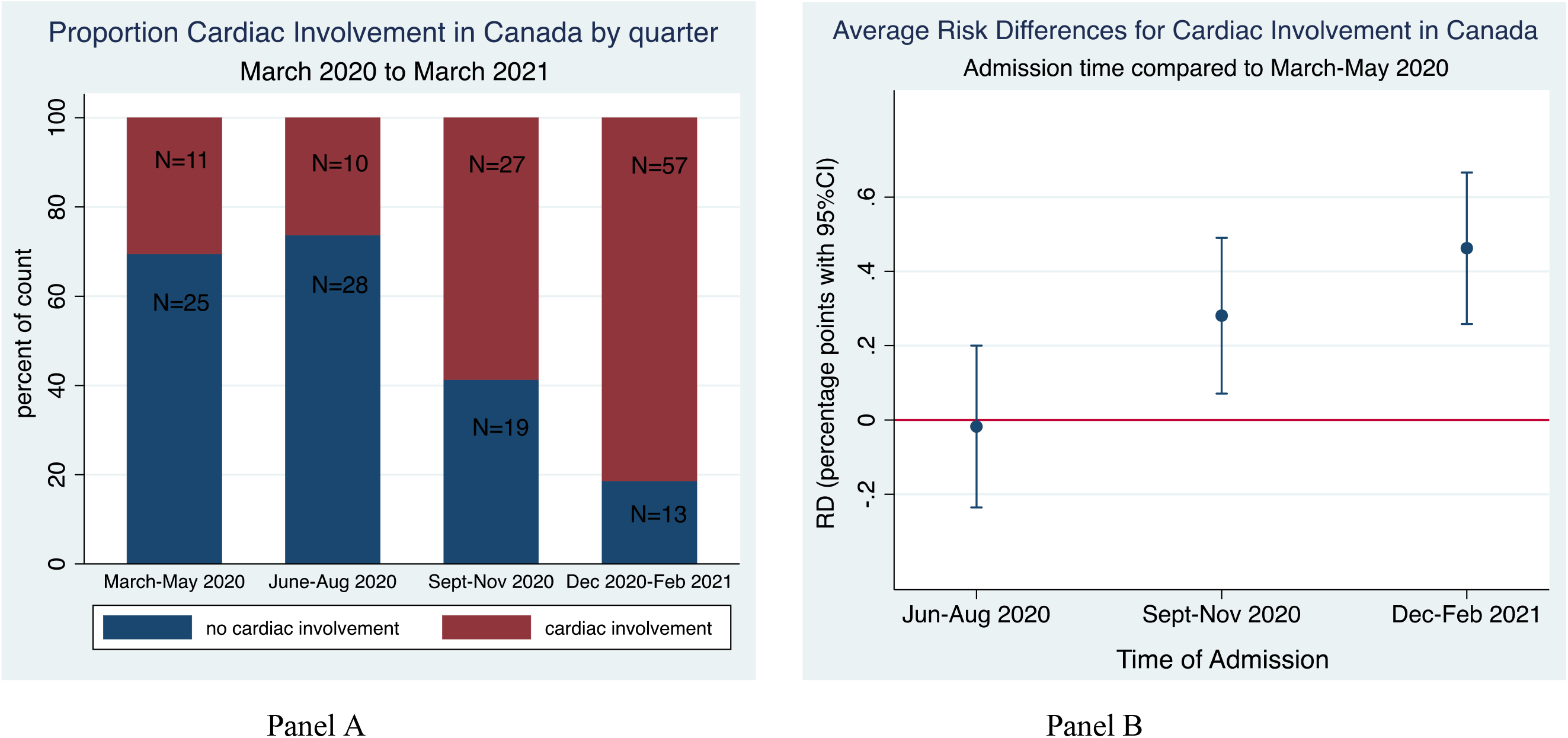
Cardiac involvement in 190 children admitted in Canada with MIS-C by quarter of the year. Cardiac involvement was defined as features of myocardial dysfunction, pericarditis, valvulitis, or coronary abnormalities (including echocardiographic findings or elevated Troponin/NT-proBNP) Panel A: N is the absolute number of cases in the group. The Y-axis reflects the proportion of total cases admitted per quarter. Panel B: Association time of admission and severe disease, defined as need for ICU admission in Canada. Average marginal effects are presented with adjusted RDs from the multivariable logistic regression. The reference used for RDs is admission to the hospital March-May 2020 with a diagnosis of MIS-C. Model is adjusted for sex, age, country, presence of comorbidity, coinfection, MIS-C laboratory confirmation.

